# The Great Genotyper: A Graph-Based Method for Population Genotyping of Small and Structural Variants

**DOI:** 10.1101/2024.07.04.24309921

**Authors:** Moustafa Shokrof, Mohamed Abuelanin, C.Titus Brown, Tamer A. Mansour

## Abstract

Long-read sequencing (LRS) enables variant calling of high-quality structural variants (SVs). Genotypers of SVs utilize these precise call sets to increase the recall and precision of genotyping in short-read sequencing (SRS) samples. With the extensive growth in availabilty of SRS datasets in recent years, we should be able to calculate accurate population allele frequencies of SV. However, reprocessing hundreds of terabytes of raw SRS data to genotype new variants is impractical for population-scale studies, a computational challenge known as the N+1 problem. Solving this computational bottleneck is necessary to analyze new SVs from the growing number of pangenomes in many species, public genomic databases, and pathogenic variant discovery studies.

To address the N+1 problem, we propose The Great Genotyper, a population genotyping workflow. Applied to a human dataset, the workflow begins by preprocessing 4.2K short-read samples of a total of 183TB raw data to create an 867GB Counting Colored De Bruijn Graph (CCDG). The Great Genotyper uses this CCDG to genotype a list of phased or unphased variants, leveraging the CCDG population information to increase both precision and recall. The Great Genotyper offers the same accuracy as the state-of-the-art genotypers with the addition of unprecedented performance. It took 100 hours to genotype 4.5M variants in the 4.2K samples using one server with 32 cores and 145GB of memory. A similar task would take months or even years using single-sample genotypers.

The Great Genotyper opens the door to new ways to study SVs. We demonstrate its application in finding pathogenic variants by calculating accurate allele frequency for novel SVs. Also, a premade index is used to create a 4K reference panel by genotyping variants from the Human Pangenome Reference Consortium (HPRC). The new reference panel allows for SV imputation from genotyping microarrays. Moreover, we genotype the GWAS catalog and merge its variants with the 4K reference panel. We show 6.2K events of high linkage between the HPRC’s SVs and nearby GWAS SNPs, which can help in interpreting the effect of these SVs on gene functions. This analysis uncovers the detailed haplotype structure of the human fibrinogen locus and revives the pathogenic association of a 28 bp insertion in the FGA gene with thromboembolic disorders.

## 2 Introduction

Maya Angelou eloquently stated, “In diversity, there is beauty and there is strength.” This principle is particularly relevant to genomics studies, emphasizing the importance of exploring genetic diversity across large cohorts and populations. Such research is crucial for advancing our understanding of evolution [1, 2], genetic adaptations [3], and gene-disease associations [4, 5]. Genetic diversity originates from various mutations, including single nucleotide variants (SNVs), small insertions and deletions (less than 50 base pairs), and structural variants (greater than 50 base pairs). Notably, structural variants (SVs) enhance genomic diversity fifteen times more than SNVs [6] and significantly affect gene function [7]. However, SVs are understudied compared to smaller variants due to the limitations of short-read sequencing (SRS), which often yields high false positive rates and inconsistent recall, varying from 10% to 70% [8]. In contrast, long-read sequencing (LRS) provides more reliable precision and recall rates [8] and is used in both mapping [9, 10, 11] and assembly-based approaches [12], the latter of which helps mitigate mapping biases to a linear genome reference. Despite its advantages, LRS remains prohibitively expensive for comprehensive population-scale analysis, and the volume of LRS data available still pales in comparison to that of SRS. As a result, there is a pressing need to develop computational techniques that utilize the precise variant discovery capabilities of LRS while maximizing the extensive data produced by SRS.

To address the shortcomings of SV callers from short-read sequencing, specialized genotypers analyze the presence and genotype of SVs, whether identified through variant calling from SRS or LRS, in SRS samples [13, 14, 15, 16, 17]. Tools such as Paragraph [14] and Graphtyper2 [16] realign reads to a variation-aware graph, minimizing mapping bias and determining genotypes from this realignment. Pangenie [17] uses k-mers specific to all potential alleles to genotype phased variants from pangenomes, minimizing mapping bias. Furthermore, Pangenie integrates genotyping and imputation, utilizing the phasing information from the pangenome to infer genotypes in regions lacking coverage, thereby achieving superior performance compared to other SV genotypers. Unlike these single-sample genotypers, muCNV utilizes population data to refine genotyping by modeling read mapping statistics across multiple samples, enhancing genotyping accuracy [15].

SV genotypers generally achieve higher recall and precision compared to direct variant calling in SRS samples. For instance, Huddleston et al. [18] used LRS to analyze SVs in two human genomes and found that 90% of these SVs were missing in the 1000 Genomes call set, yet 61% could still be genotyped using SRS. Recent population-scale studies have therefore adopted a combined approach of variant calling and genotyping: initially, variants are identified from a few LRS samples or numerous SRS samples, and then the identified SVs are merged and genotyped in a larger SRS cohort [19]. For instance, Kirsche et al. [20] used Paragraph [14] to genotype variants from 31 LRS samples in a cohort of 1.3k SRS samples from the 1000 Genome Project (1kGP) [21]. Similarly, Graphtyper2 was employed to build graphs from SVs detected in 50k Icelandic SRS samples [16] or 2k dog SRS samples [22], which were then re-genotyped using the same SRS samples to improve recall. With the same concept in mind, the Human Pangenome Reference Consortium (HPRC) [23] applied Pangenie to genotype the pangenome variants in 3.2k SRS samples from 1kGP [21]. Similarly, Goo Jun et al. [59] used MuCNV to jointly genotype TopMed SVs in 139k SRS samples. These genotypers enable large-scale population genotyping of gene catalogs, pangenomes, and candidate disease-associating variants.

The current SV genotypers, while fast and scalable, face significant challenges at the population level. These genotypers require downloading and reprocessing all the raw SRS data to genotype even a single new variant, a demand that is increasingly impractical. This issue exemplifies a computational challenge known as the N+1 problem [67]. In today’s era of extensive sequencing, new lists of variants emerge daily, and a reliable estimation of their allele frequencies is important for interpretation. For instance, the number of pangenomes for humans [23, 24] as well as numerous other species [25, 26, 27, 28] is increasing. Similarly, databases like dbVar [29], genomeAD [60], TopMed [30], and ClinVar [31] are constantly expanding their variant collections. The N+1 challenge also affects disease gene discovery studies in probands [32]. LRS can produce phased, high-quality SVs, and identifying pathogenic variants involves filtering out common variants and focusing on rare ones. However, matching these variants in public databases poses challenges, and the reliability of allele frequencies in SV catalogs is dubious when calculated in small or distinct subpopulations or when using methods with low recall. Therefore, solving this computational bottleneck is crucial to optimize the usage of genomic data for advancing precision medicine and enhancing our understanding of genetic diversity.

We therefore introduce “The Great Genotyper,” an alignment-free population genotyping pipeline for both structural and small variants. This pipeline can accurately genotype four thousand human WGS samples in just hours, bypassing the need for 183TB of raw sequence data. Instead, it utilizes an 867GB Counting Colored Debruijn Graph (CCDG), which is constructed once from the population’s raw sequences by extracting k-mers and their counts. The CCDG provides a viable solution to the N+1 problem as it can be reused to genotype any new variant list for this population. In addition, it utilizes population-derived information to improve the quality of genotyping, creating an imputation panel for SVs using a pangenome, and annotating SVs by their linkage to nearby GWAS SNPs.

## 3 Result

### 3.1 The Great Genotyper: A Workflow for Genotyping Small and Structural Variants in Thousands of Short-Read Samples

The Great Genotyper solves the N+1 problem by deploying two independent workflows. The first is an indexing workflow that creates a CCDG using SRS to represent the population (Figure 1A). Once created, the CCDG can be reused by a population genotyping workflow (Figure 1B) to genotype a pangenome, phased variants, or unphased variants in the cohort of SRS samples.

**Figure 1:**
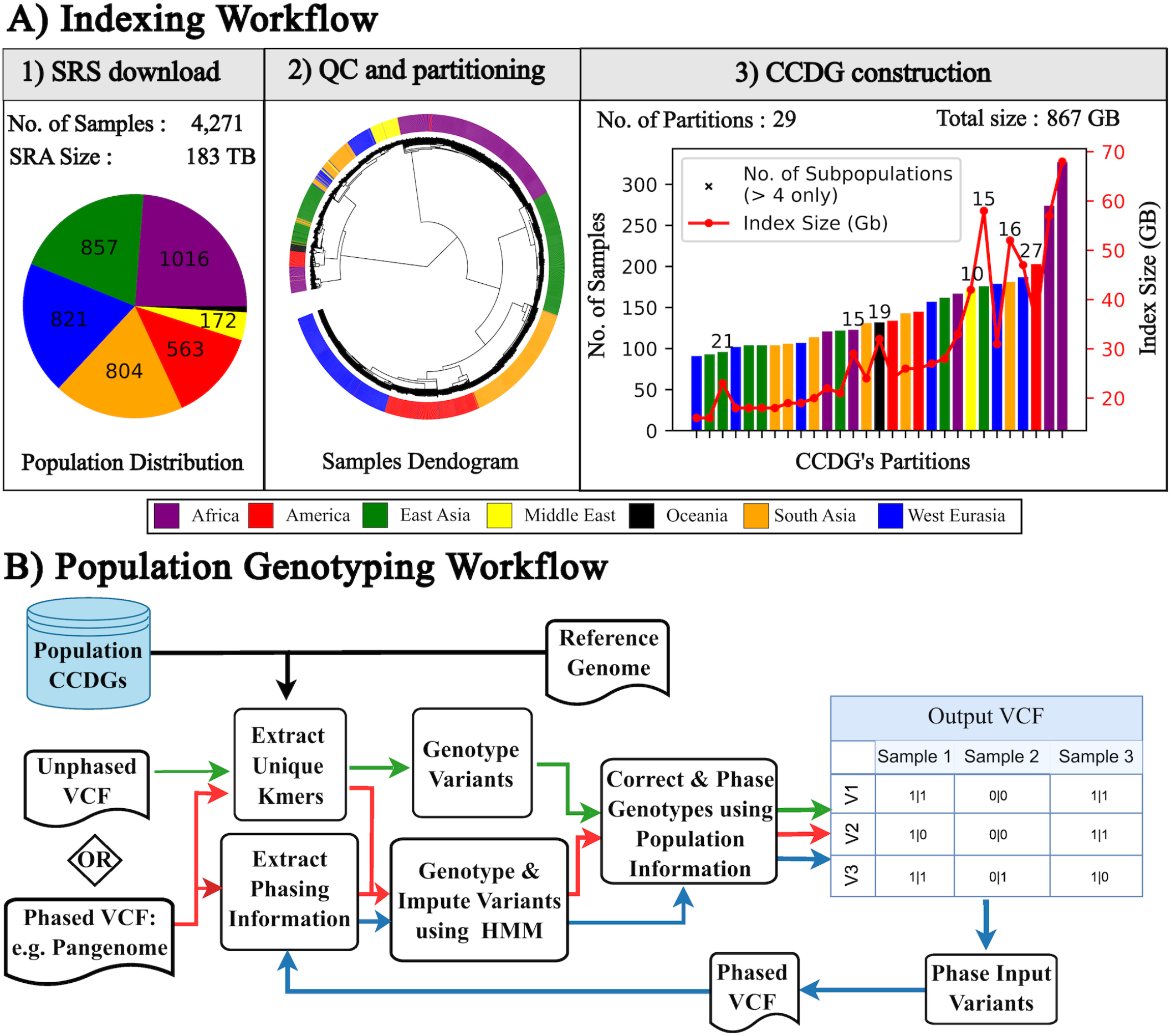
The Great Genotyper workflows. The indexing workflow (A) depicts the high-level pipeline for creating Population CCDGs. The workflow downloads and computes the unitigs of each sample individually (A1). A sourmash signature is calculated for each sample to be used for alignment-free quality control and sample partitioning (A2). Lastly, a subgraph is created for each partition of samples (A3). The genotyping workflow (B) describes three population genotyping workflows illustrated with a different color of arrows: The HMM workflow (red) genotypes and imputes phased variants using a high-quality HMM model, the k-mer-based workflow (green) rapidly genotypes unphased variants, and the two-pass workflow (blue) enhances the recall of the k-mer-based workflow by genotyping its output phased variants using the HMM workflow.

For human populations, we download 4.2K high-coverage (30x) whole genome sequencing (WGS) samples from three projects: 1KGP [21], HGDP [33], and SGDP [34]. This data represents 140 populations worldwide (Figure 1A and Supplementary Figure 1). While CCDGs are much smaller than raw sequences, creating a single CCDG for thousands of samples would result in a complex and memory-intensive structure. Therefore, the indexing workflow partitions closely related samples into separate groups, creating individual CCDGs for each (Figure 1A.2). This workflow starts by downsampling raw sequences into representative summaries (i.e. FracMinHash sketches calculated by sourmash [66]. These sketches are used for quality control including estimate calculation of sequencing depth, genome coverage, possible contamination, and sex prediction [61]. For example, this analysis reveals samples with unexpectedly low coverage of the human genome as well as four discrepancies from the sex provided in the metadata (Supplementary Figures 2 and 3). After that, a pairwise comparison of sourmash signatures enables the creation of a dendrogram (Figure 1A.2). Based on this dendrogram, 29 partitions of closely related samples are identified (Figure 1A.3). Finally, Metagraph [62] is used to create a CCDG for each partition, resulting in individual files ranging from 16-68 GB with a total size of 867 GB.

**Figure 2:**
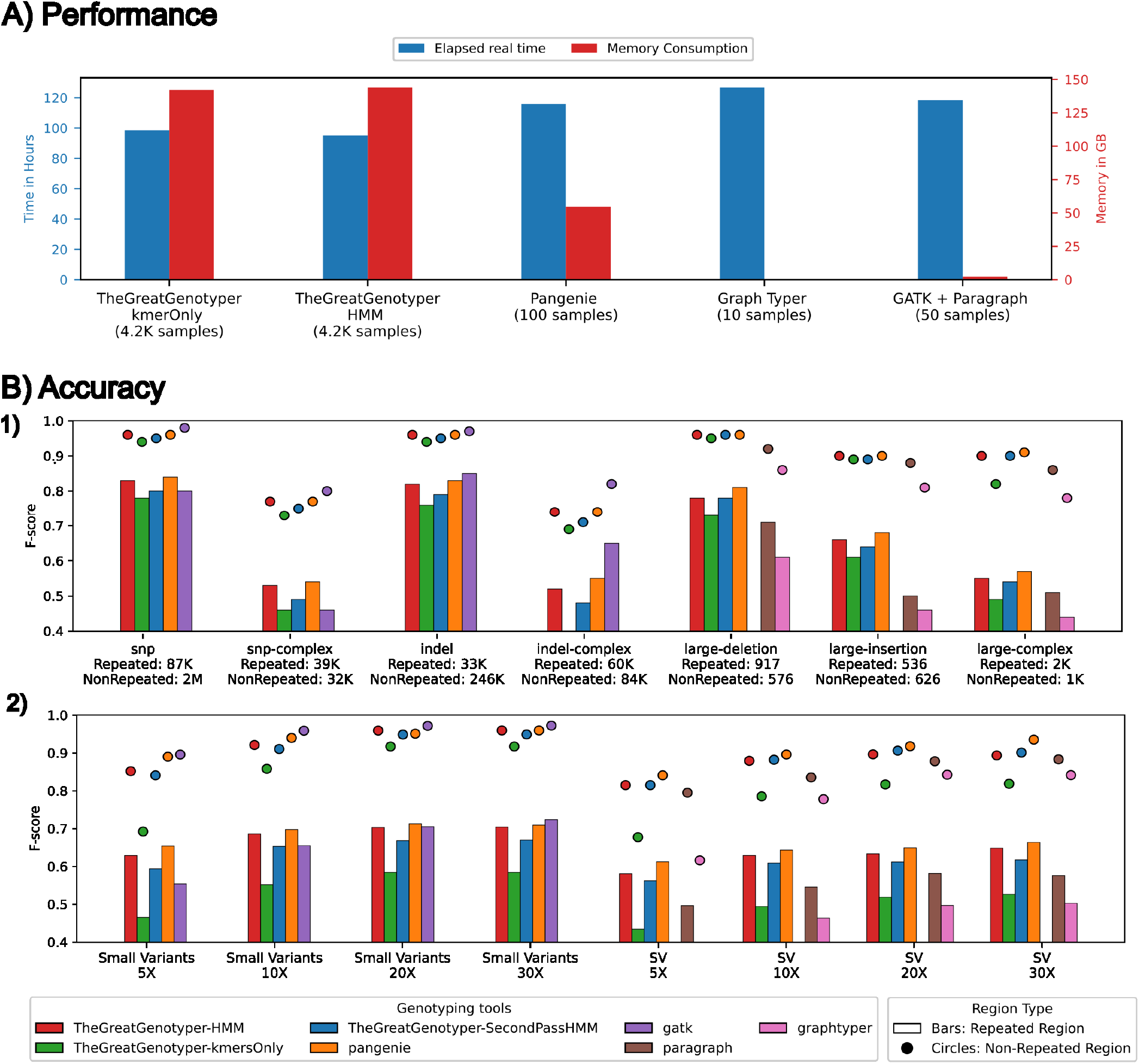
The Great Genotyper provides unparalleled performance compared to the state of the art, with no compromise on accuracy. **Panel A** shows the running time and memory usage of different tools used to genotype 4.5 million phased variants (including structural variants and small variants). The Great Genotyper is currently genotyping 4,200 samples at 30x coverage, while the other genotypers are handling 10 to 100 samples. **Panel B1** illustrates the F-scores of different genotyping methods for different classes of variants. **Panel B2** illustrates the effect of coverage on the F-scores of different genotyping methods for small and structural variants. In both Panels B1 and B2, the variants are categorized based on the complexity of the genomic loci into variants located in repeated (shown as bars) and non-repeated regions (shown as circles)

**Figure 3:**
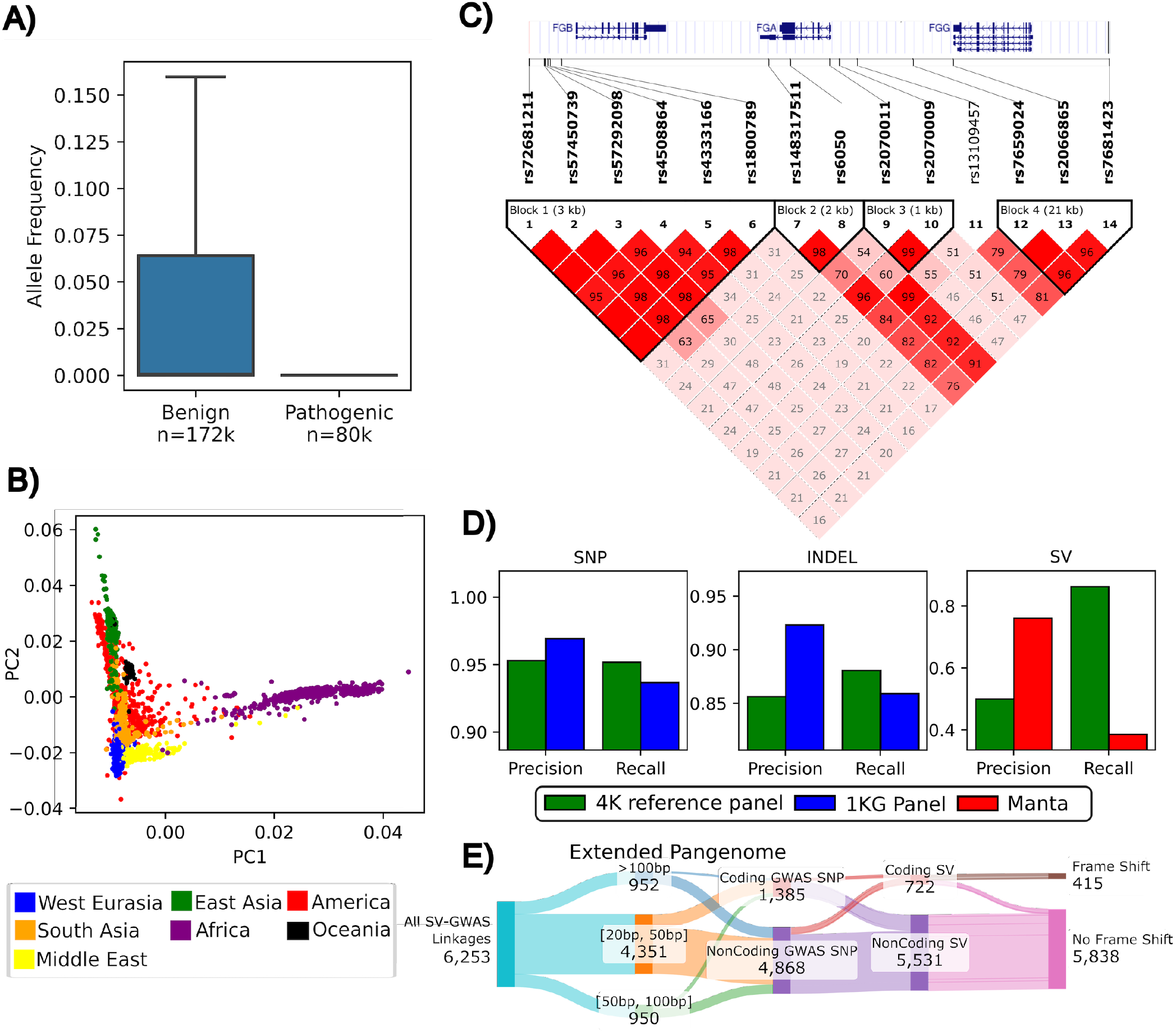
Applications of The Great Genotyper: We used the Great Genotyper to genotype all ClinVar and HPRC pangenome variants in 4k human samples. **Panel A** is a box plot of the distinctive distributions of population allele frequencies for ClinVar variants when stratified by the pathogenicity of the variants (outliers are not displayed). **Panel B** is a plot of the first two principal components from a PCA for the genotypes of the HPRC pangenome variants; the 4k samples are colored by their ancestry. **Panel C** is an LD heatmap that highlights the associations of an insertion (dbSNP: rs148317511) and multiple GWAS SNPs including rs6050-C; a peak associating SNP in a GWAS study of the circulating fibrinogen. **Panel D** shows the precision and recall of small and structural variant imputation using the 4k reference panel in comparison to small variant imputation using the 1000 Genome panel and calling SVs using Manta. **Panel E** presents a Sankey plot summarizing 6.2K linkage associations between SVs from the HPRC pangenome and the GWAS catalog. The columns stratify linkages based on various traits of both SVs and GWAS: SV size, GWAS SNP impact on coding regions, SV impact on coding regions, and SV-induced frameshifts.

Building upon the CCDGs created in the indexing workflow, the genotyping workflow (Figure 1B) empowers the analysis of any variant list across all samples without requiring raw reads or mapping. It begins with three key inputs: a list of pre-generated CCDGs, a reference genome and a variant list (phased or unphased). Depending on needs, three different workflows can be chosen: A) k-mer-based workflow: Efficiently genotypes unphased variants. B) Hidden Markov model (HMM) workflow: Handles both genotyping and imputation for phased variants. C) Two-pass workflow: Genotypes and imputes unphased variants, leveraging population information to determine their phase and impute missing data.

Both the k-mer-based and HMM workflows start by extracting k-mers unique to the variant regions and querying their count data for all samples within the CCDGs (Figure 1B). The k-mer-based workflow determines initial genotypes by comparing the counts of unique k-mers to the average sample coverage for each sample. This identifies variants present in each sample without relying on phasing information. In contrast, the HMM workflow tackles phased variants by genotyping and imputing them using the Hidden Markov Model (HMM) implemented in Pangenie [17]. This enables the imputation of genotypes in regions with low coverage or complexity. Following initial genotyping, both workflows undergo a two-step refinement. The first step is to filter low-quality genotypes after comparing the genotype qualities for each variant across all samples. The second step utilizes Beagle [35, 36] to statistically impute low-confidence genotypes and phase the resulting variants.

The third workflow is a pipeline to genotype and impute unphased variants. It starts by running the k-mer-based workflow to create a reference panel using the input variants and samples in the CCDGs. This reference panel is then used to phase the input variants. After that, the HMM workflow is employed on the phased variants to obtain more precise genotypes in indexed population.

### 3.2 Achieving Population Genotyping in a Matter of Hours with no Decrease in Accuracy

The performance of the Great Genotyper was evaluated for the k-mer-based workflow (for unphased variants) and HMM workflow (for phased variants). The Great Genotyper could genotype 4.5 million variants across 4.2K WGS samples in approximately 100 hours, utilizing 32 cores and 145 GB of memory, as depicted in Figure 2A. To put this performance into context, Pangenie and GraphTyper2 required nearly an hour and 12 hours, respectively, to genotype the same 4.5 million variants in a single sample using the same machine. Extrapolating this duration, Pangenie would take approximately six months to complete genotyping of the same dataset, while GraphTyper2 needs six years to finish. This underscores the efficiency of the Great Genotyper in both operational modes.

For benchmarking of precision and recall of the Great Genotyper with other state-of-the-art genotypers, we genotyped SVs and small variants derived from the NA12878 haploid-resolved assemblies using the 30x SRS of HG00731 (See Methods and Supplementary Figure 4 for the design of benchmarking and Figure 2 for the detailed results).

The Great Genotyper’s HMM and Pangenie exhibit superior F-scores for phased SVs, achieving 0.91 in non-repetitive regions. Paragraph and the k-mer-based workflow follow closely with F-scores of 0.88 and 0.87 for unphased SVs. Intriguingly, the two-pass workflow accurately predicts the phasing information, boosting the F-score back to 0.91. In contrast, Graphtyper trails with an F-score of 0.80. The challenges increase in repetitive regions, where variability in results is more pronounced. Here, Pangenie and the HMM workflow score 0.63 and 0.61, respectively, followed by Paragraph and the k-mer-based workflow at 0.55. However, the two-pass workflow enhances the k-mer-based approach’s F-score to 0.6, while Graphtyper lags with an F-score of 0.48.

For small variants, GATK leads, achieving F-scores of 0.97 and 0.70 in non-repetitive and repetitive regions, respectively. The Great Genotyper’s HMM and Pangenie are close behind with F-scores of 0.95 in non-repetitive areas. The k-mer-based workflow scores 0.93, improving slightly to 0.94 with the two-pass workflow. In repetitive regions, Pangenie matches GATK’s 0.70 F-score, while the HMM workflow slightly trails at 0.69. The k-mer-based workflow struggles in these regions and scores 0.6 but is improved to 0.65 by the two-pass workflow. Overall, the Great Genotyper consistently demonstrates competitive genotyping accuracy compared to Pangenie across most scenarios, and it represents the most accurate option for genotyping unphased SVs with the two-pass workflow.

Sequencing depth impacts the genotyping accuracy, as depicted in Figure 2B2. Notably, all genotypers exhibit reduced accuracy at sequencing depths of 10x and 5x. Genotypers that incorporate phasing information, such as The Great Genotyper’s HMM and two-pass workflows, as well as Pangenie, show the smallest decrease in accuracy. For instance, the accuracy of SV genotyping by The Great Genotyper’s HMM and Pangenie at 5x coverage drops by 8% and 9% in non-repetitive regions, and 7% and 5% in repetitive regions, respectively. The k-mer-based workflow experiences a decrease of 14% and 9%, which the two-pass model returns to 7% and 5% in non-repetitive and repetitive regions, respectively. Last, Graphtyper’s accuracy diminishes by 12% and 22% in non-repetitive and repetitive regions, respectively.

The reduction in sequencing depth from 30x to 5x similarly affects the accuracy of small variant genotyping in both non-repetitive and repetitive regions. Pangenie exhibits the smallest accuracy decline, by 7% and 5%, followed by The Great Genotyper’s HMM with 11% and 7%, and GATK with 8% and 17%. The k-mer-only model suffers a significant drop of 22% and 11%, but this is mitigated by the two-pass model to 10% and 7% in non-repetitive and repetitive regions, respectively.

### 3.3 Facilitating Population Studies for Small and Structural Variants

#### 3.3.1 The Great Genotyper can help to find pathogenic variants

Filtering common variants is a widely used strategy in disease association studies. ClinVar, a public database, catalogs genomic variations in humans and their impact on health [31]. As a proof of concept, the k-mer-based workflow is applied to genotype the ClinVar database variants in the 4k samples of the CCDG index. Consistent with expectations, almost all pathogenic variants exhibit zero allele frequency in this healthy population, whereas benign variants display a broader range of frequencies (Figure 3A). This demonstrates that calculating allele frequencies for a list of suspected variants in this indexed cohort is a reliable metric for prioritizing rare variants in studies of their pathogenic potential.

#### 3.3.2 Generation of 4k reference panel by Genotyping HPRC Variants in 4K Samples

The current HPRC pangenome comprises 88 haplotypes (Citation). As previously described [23], decomposing the pangenome yields a phased VCF containing 26.8 M variants (See Supplementary Table 1 for the summary count per variant type). The HMM workflow is used to genotype these variants in the prebuilt CCDG. The resulting output is a phased VCF of the HPRC variants in the indexed 4K samples, creating a new 4K reference panel. Principal Component Analysis (PCA) on the genetic variation within this 4k reference panel confirms the expected distribution of populations studied in the 1kGP, paving the way to generate cost-efficient similar panels for several other species (Figure 3B). Subsequent sections will explore how this panel can facilitate various genomic applications.

#### 3.3.3 Impute SV by using the 4k reference panel

Genotype imputation is a statistical method that predicts unobserved genotypes using reference sequences, thereby enhancing the density and scope of genetic analyses at reduced costs. This technique is especially valuable in increasing the power and consistency of genetic studies, including genome-wide association studies (GWAS) and fine-mapping efforts [37]. The 4k reference panel may replace the panel generated by the 1kGP project [21] while enabling the imputation of structural variants (SVs). In this section, we demonstrate the precision and recall of imputing both small and structural variants using the 4k reference panel. Initially, pseudo-microarray variant calls are generated using the HG002 sample from the Genome in a Bottle (GIAB) project [38] by extracting variants at sites used in the Illumina Infinium OmniExpress-24, simulating microarray genotyping. The 4k reference panel is then employed to impute both small and structural variants. For benchmarking purposes, the 1kGP reference panel is used exclusively for imputing small variants, with no similar panel available for SVs. Instead, SV calling from 30x SRS using Manta serves as an alternative. The output VCFs are compared against gold standard GIAB datasets using hap.py (v0.3.12) [39] for small variants, and truvari (v3.5.0) [63] for SVs. The 4k reference panel exhibits commendable precision and recall for the imputation of both types of variants, as depicted in figure 3D. When compared to the 1kG reference panel, it displayed some reduced precision compensated by an increase in recall for SNPs and indel imputation. Conversely, the 4k reference panel shows remarkable results in imputing common SVs, achieving an impressive recall of 86%. This imputation recall surpasses the recall of SV calling from 30x SRS using Manta. These results highlight how the 4k reference panel can be leveraged to augment microarray genotypes with common SVs.

#### 3.3.4 Fine Mapping of GWAS SNPS using SVs from the 4k reference panel

The 4k reference panel provides detailed insights into the structure of common haplotypes composed of small and structural variants. In particular, it allows the exploration of linkage disequilibrium (LD) between SVs and neighboring variants known to be associated with phenotypic changes. We initiate our investigation by annotating the SVs in the 4k reference panel using AnnotSV (v3.3.6) [40]. This reveals that approximately 463K SVs affect gene structures. Proceeding further, we compute the pairwise LD for each of these variants with all the variants located within a 1MB window surrounding them. Our analysis indicates that 91K SVs exhibit a strong association with a neighboring variant, having an r2 value greater than 0.8.

We utilize the identified associations to illuminate potential causal variants in GWAS studies. Among the 91K SVs, 3,744 are found in strong linkage with GWAS SNPs. We compiled a table that includes these SVs, their annotations, associated GWAS SNPs, and other relevant metadata (see the ‘Data and Code Availability’ section). This table should be a valuable resource elucidating the phenotypic effects of common SVs and help pinpoint some causal variants of the traits examined in these GWAS studies. Figure 3E summarizes of the associations found in the table. Notably, 722 of these SVs impact the coding regions of genes, with 415 causing frameshift mutations.

We explore a specific example from our list in figure 3C, focusing on the Human fibrinogen locus on chromosome 4. This 50-kilobase region includes three fibrinogen genes: the central FGA gene encodes the alpha chain, flanked by FGB and FGG encoding the beta and gamma chains, respectively [41]. Our reference panel shows an insertion of 28 bp at chr4:154584089 (dbSNP: rs148317511; ClinVar: RCV000247066) in a high linkage (r2=0.98) with rs6050-C; a missense mutation in FGA associating with venous thromboembolism [42, 43, 44, 45, 44] and chronic thromboembolic pulmonary hypertension [45, 46]. The insertion is reported in ClinVar as a benign variant. Surprisingly, further digging in the literature shows that the variant was once known as the Taq I polymorphism because it created an additional restriction site for Taq I [47]. The allele was found to enhance the stability of FGA mRNA in vitro [43]. This was explained by the ability of the insertion to oppose the suppressive effect of has-miR-759 on the 3 UTR of FGA [46]. These findings suggest that the ClinVar information on the variant should be revised.

Interestingly, our panel is able to capture the haplotype structure of the fibrinogen locus and shows how the 28bp insertion fits in. For example, rs6050 is known to be in high linkage with rs7681423; SNP upstream to FGG and a peak of association with *γ*^*′*^ Fibrinogen. Both SNPs are known to have no significant association with total fibrinogen levels and no linkage with rs1800789; SNP in FGB shows the strongest association with total fibrinogen level, but not with *γ*^*′*^ fibrinogen [48]. The panel confirms these relationships between the three SNPs and shows that the insertion allele has some linkages (r2=76) to rs7681423 and no linkage to rs1800789. Also, the panel shows a unique haplotype (r2=96) of the insertion and rs2070011-A; an allele of CFA’s promoter causing higher expression of the gene. This haplotype is different from the haplotype of rs6050 and rs7681423.

## 4 Discussion

The Great Genotyper serves as a practical solution for population genotyping at massive scales. It provides the ability to genotype a new set of variants, whether small or structural, in thousands of SRS samples in just a matter of hours. More importantly, it provides a novel solution for the chronic N+l problem by eliminating the need to download and process terabytes of raw sequencing data. Instead, the Great Genotyper operates using a prebuilt CCDG, effectively decoupling intensive data preprocessing from the actual genotyping process. In terms of input, the Great Genotyper is versatile; it accepts any set of phased or unphased variants, along with the reference genome. The outcome is the phased genotypes of all input variants in the indexed samples. In this manuscript, 183 TB of SRA files for 4K human SRS samples are indexed to generate an 867 GB CCDG to enable unprecedented efficiency in calculating allele frequencies of any list variants in the human population. As a proof of concept, the index is used to genotype the HPRC pangenome variants as an example for phased variants as well as genotyping all unphased ClinVar variants.

The Great Genotyper does not sacrifice quality for scalability. On the contrary, the scalability empowers the Great Genotyper to jointly genotype thousands of samples, which, in turn, enhances the genotyping quality even more. K-mer-based genotypers such as Nebula and Pangenie have previously demonstrated the potential of k-mers for precise genotyping. They leverage the specificity of variant-specific k-mers, using shifts in the counts of these k-mers as indicators to genotype the variants. The Great Genotyper reinforces this approach, considering the counts of these k-mers across an entire population of samples. This innovation facilitates the calculation of a confidence measure for each genotype based on the collective population data. Furthermore, the tool is equipped to impute missed genotypes through a two-tiered approach. Initially, imputation is rooted in the phasing information of the variants, either provided as input or derived from the large cohort genotypes. Subsequently, the Great Genotyper integrates Beagle, leveraging the high-confidence genotypes within the population to further impute genotypes. This dual-phase imputation process ensures that the Great Genotyper can deliver performance on par with Pangenie, even if some data is compromised during the k-mer count preprocessing while indexing to enable better data compression as described in Supplementary Figure 7.

The enhanced accuracy and scalability of the Great Genotyper paves the way for valuable downstream applications in genomics. For instance, accurate allele frequencies can now be directly derived from sequences rather than merging information from sparse studies or variation databases that rely on variant calling in SRS studies. Such accurate determination of allele frequencies can play a pivotal role in pinpointing causal variants in disease-gene discovery studies. Furthermore, simultaneous genotyping and phasing of common variants enables dramatically improved resolution for understanding the haplotype structure within and across populations. As an example, genotyping the HPRC pangenome variants in 4k samples produces what we call “the 4k reference panel (4kRP)”. We show how the 4kRP can be used to impute common SVs with a recall rate that surpasses some short-read callers like Manta.

Taking our analysis further, we explore the 4kRP for SVs in high LD with known GWAS SNPs. We limit our focus to 91K SV variants impacting gene structures. Intriguingly, we discover that approximately half of these SVs exhibit strong associations with at least one GWAS SNP. We are optimistic that our findings will contribute to a deeper comprehension of the relationship between genotype and phenotype concerning these structural variants.

Although the Great Genotyper is effective in generating high-quality genotypes for both small and structural variants, it does have certain limitations. First, some variants cannot produce specific k-mers because the k-mers from the alternate sequences may also be present in other parts of the genome. Such variants cannot be genotyped precisely by k-mer-based approaches. This limitation, however, is partially offset through imputation. Furthermore, genotyping copy number variants is beyond the capabilities of the current version of the Great Genotyper. While it is not an insurmountable challenge, it requires development of a dedicated genotyping model. Another constraint is that the Great Genotyper utilizes two separate imputation models, as they are implemented in two distinct tools, Pangenie and Beagle. A unified model tailored specifically for imputing genotypes using the k-mers in the CCDG could both enhance the accuracy as well as boost performance.

The Great Genotyper opens many doors for future genomic applications. Creating more CCDGs to represent specific subpopulations or individuals exhibiting specific traits, like autism, is crucial for understanding the role of genomics in these cohorts. Moreover, while most population studies have been conducted on humans [49], this approach is applicable to man y other organisms. The Sequence Read Archive (SRA) [50] is a vast reservoir of short-read samples for non-human organisms. Generating CCDGs for these samples will facilitate population-scale studies for other species.

The current CCDG for the human population, and the additional CCDGs to be created for other cohorts, are invaluable resources with potential applications that extend beyond genotyping. For instance, variants can be directly called from the graph using methods such as Corticall [51]. Additionally, it can aid in subsetting pangenomes by selecting segments of the pangenome that have k-mers present in a specific population, thereby creating a more streamlined pangenome tailored to that population. We encourage the community to explore and uncover more ways to harness the extensive genomic diversity revealed by the CCDG.

## 5 Conclusion

The Great Genotyper can transform population genotyping into a routine task using a flexible CCDG representation of populations. Its scalability allows the improvement of genotyping quality by using population information. The tool’s practicality aids in expanding variant lists into broader dimensions, revealing complex genomic details. We demonstrate its potential in applications such as creating SV imputation panels, finding SV associations with variants from databases like the GWAS catalog, and accurately calculating population allele frequencies. The CCDG, comprising 4,000 human samples, contains a vast genomic variation spectrum, accessible through The Great Genotyper or other methods, leading to enhanced genomic insights. Producing more CCDGs for additional cohorts or species will further optimize the use of existing SRS samples.

## 6 Data and Code Availability

The code for The Great Genotyper is publicly available on GitHub at the following URL: https://github.com/dib-lab/TheGreatGenotyper. The benchmarking code used in our study can also be found on GitHub at this URL: https://github.com/dib-lab/TheGreatGenotyper_benchmark. The indexes used in our project are hosted on our server and can be accessed at this URL: https://farm.cse.ucdavis.edu/∼tahmed/GG_index/. We have also provided several use cases which can be found at this URL: https://github.com/dib-lab/TheGreatGenotyper_usecases. The genotyped pangenomes are available at this URL: https://farm.cse.ucdavis.edu/∼mshokrof/4k_reference_panel/. The LD list and the GWAS SV Associations can be found at this URL: https://farm.cse.ucdavis.edu/∼mshokrof/GWAS_associations/. Lastly, the ClinVar genotyped data can be accessed at this URL: https://farm.cse.ucdavis.edu/∼mshokrof/The_great_genotyper_clinvar/.

## 7 Methods

### 7.1 Short Read Samples Preprocessing and Clustering

Upon the download of each sample, kmc [52] was used for k-mer counting with a minimum count of 3 to filter out singletons and doubletons, which are likely sequencing errors. In addition, Metagraph [62] was utilized to identify the unitigs and retain only the average k-mer count per unitig, thus smoothing k-mer counts. This smoothing reduced the size of the k-mer counts to one-tenth while maintaining high genotyping accuracy (see below). Subsequently, alignment-free quality control was done using Snipe [61]. In brief, a sourmash sketch was created for each sample using a k size of 51 and a subsampling scale of 10k, which entails keeping a single hash for every 10,000 k-mers. A similar sketch at the same scale was created for the GRCh38 reference genome. Intersection of both signatures enabled approximate estimation of the genome coverage and sequencing depth as well as sex confirmation (Supplementary Figures 2 and 3). Subsequently, kSpider [64] calculated pairwise similarities between all samples based on their sourmash sketches. To alleviate skew from the sex chromosomes, the sketch of chrY was subtracted from all samples. Hierarchical clustering was employed using the Scipy library [65] to construct a dendrogram visualized in Supplementary Figure 6 by iTOL [53]. From the dendrogram, twenty-nine clusters were extracted and subsequently refined manually to ensure each encompasses between 100-350 samples.

### 7.2 Determining The Best Indexing Parameters

We investigated the influence of sample preprocessing on genotyping accuracy to determine the best parameters for optimal results. Multiple CCDGs were generated from sub-samples of the HG00731 SRS at sequencing depths of 5x, 10x, 20x, and 30x. Each CCDG was constructed using a different set of parameters, which are summarized in Supplementary Table 2, along with the final sizes of the CCDGs. Benchmarking was done as described in Supplementary Figure 4 and later in the methods.

Results in Supplementary Figure 7 and Table 2 indicate that preprocessing methods do not impact samples with coverage exceeding 20x. For coverages of 10x and 5x, logging the counts is the most influential, significantly decreasing both the F-score and the final CCDG size. On the other hand, smoothing leads to a nominal drop in the F-score but notably reduces the CCDG size. Cleaning had a moderate impact on the F-score and caused a slight reduction in the CCDG size. These findings are instrumental in guiding our final decision to use smoothing of k-mer counts as the only preprocessing for input samples.

### 7.3 Genotyping Workflow

The Great Genotyper adopts the genotyping model from Pangenie [17] to implement two genotyping workflows; one for genotyping unphased variants using k-mer counts, and another for genotyping and imputing phased variants using k-mer counts and phasing information. For the phased variants, the Great Genotyper employs the Pangenine HMM model, which is based on the Li-Stephen model [54]. For the unphased variants, we rely solely on emission probabilities to determine the most probable genotype for each variant. The genotyping models yield a confidence measure for the output genotypes, deducing the likelihoods of all possible genotypes and selecting the one with the highest probability. The confidence level is derived from the difference between the highest probability and the other probabilities. Therefore, a larger difference correlates to higher confidence.

The components driving these confidence probabilities can primarily be distilled into two factors: the number of unique k-mers discovered for each variant haplotype and the count of these k-mers in the sample. The first factor is a constant across all samples since it is determined only from the reference genome and the variant to be genotyped. However, the second factor varies per sample. Some samples may present robust evidence for a particular genotype, while others may not due to either low coverage of the region in the sample or the exhibition of a different haplotype not present in the input haplotypes. The Great Genotyper leverages the power of having a large population in the CCDGS to filter low-quality genotypes. In this step, the median of genotype confidences is calculated for each genotype then the genotypes falling below this median are discarded. This approach allows the Great Genotyper to establish a variable threshold calculated using the results from all the samples, providing a balanced way to sift through the variants. For variants abundant in unique k-mers, this threshold will be high, while more challenging variants will have a lower threshold, accommodating the varying levels of confidence in different scenarios. The final output of this step is a reference panel comprised of the high-confidence genotypes.

Finally, Beagle [35, 36] is employed to statistically impute the filtered, low confidence genotypes using this reference panel, simultaneously phasing the resultant variants, thereby yielding phased genotypes for all samples. It is crucial to note that Beagle employs a different HMM model, albeit very similar to the one used in the HMM workflow. In Beagle, linkage disequilibrium is computed statistically from the high-confidence genotypes within the created reference panel. In contrast, the model in the HMM workflow utilizes the phasing information provided by the user in the input variants. The synergy between these two imputation methods does not only enhance the results of genotyping but also broadens the application scope for the higher quality HMM model, enabling its usage when phasing information is absent in the input VCF, as described in the two-pass workflow in Figure 1B.

### 7.4 Benchmark Experiment Design

This section outlines the experimental design for the benchmarking experiments conducted to compare the accuracy of The Great Genotyper with the state-of-the-art genotyping tools: Pangenie, GraphTyper2, Paragraph, and GATK as described previously [17]. This experiment is structured into two components. The first component involves creating a truth variant set and a query variant set.

The truth set comprised of small and structural variants is created by aligning each haplotype-resolved assembly of HG00731 against the reference genome using minimap2 [55] and identifying the variants using PAV tools [56]. The resulting two VCFs are merged using bcftools [57]. The analysis is confined to a confidence region on the reference genome, ensuring that only one segment from the assembly maps to it to avert the regions difficult to assemble. Variants within this confidence region are deemed suitable to represent the full truth in these regions. Similarly, the query VCF is created from the haplotype-resolved assemblies of NA12878 downloaded from GIAB [58]. The shared variants between the truth and query sets are categorized as true positives while those found only in the query set as true negatives.

The second component is running the genotypers and benchmarking them. Subsequently, different genotypers are executed on the query variants obtained from NA12878 and the SRS from the HG00731 sample. The genotyping results are then compared against the truth set. The benchmarking outcomes are stratified based on whether the variant is located in a repeat region or not and are also classified by type and size: SNP, Indel(*<* 50bp), Insertions/deletions(*>* 50bp), and complex Insertions/Deletions, where “complex” denotes that the variation generates more than one breakpoint.

## Data Availability

All data produced in the present work are contained in the manuscript

## 8 Author Contributions

M.S. and T.M. conceptualized the study, interpreted the results, and wrote the main draft. M.S. was responsible for the implementation of the software. T.M. supervised the work and participated in the data analysis. M.A. contributed to the experiment on reference-free QC. T.B. provided valuable feedback on the study design and reviewed the manuscript.

