## Supplementary Materials for "The Great Genotyper: A Graph-Based Method for Population Genotyping of Small and Structural Variants"

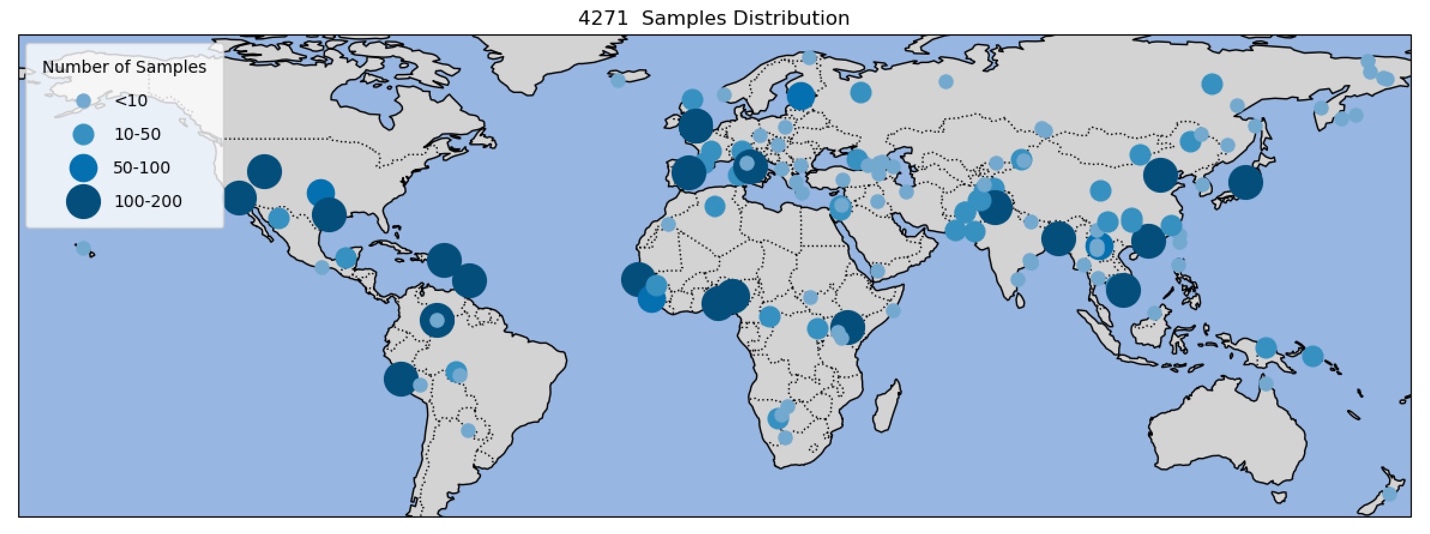

**Supplementary Figure 1:** **Global Distribution of Samples**: The map demonstrates the representational breadth of selected samples across global populations. Small circles denote samples from the SGDP and HGDP datasets, while the larger circles represent those from the 1KG project, providing an overview of the coverage of world populations by these samples.

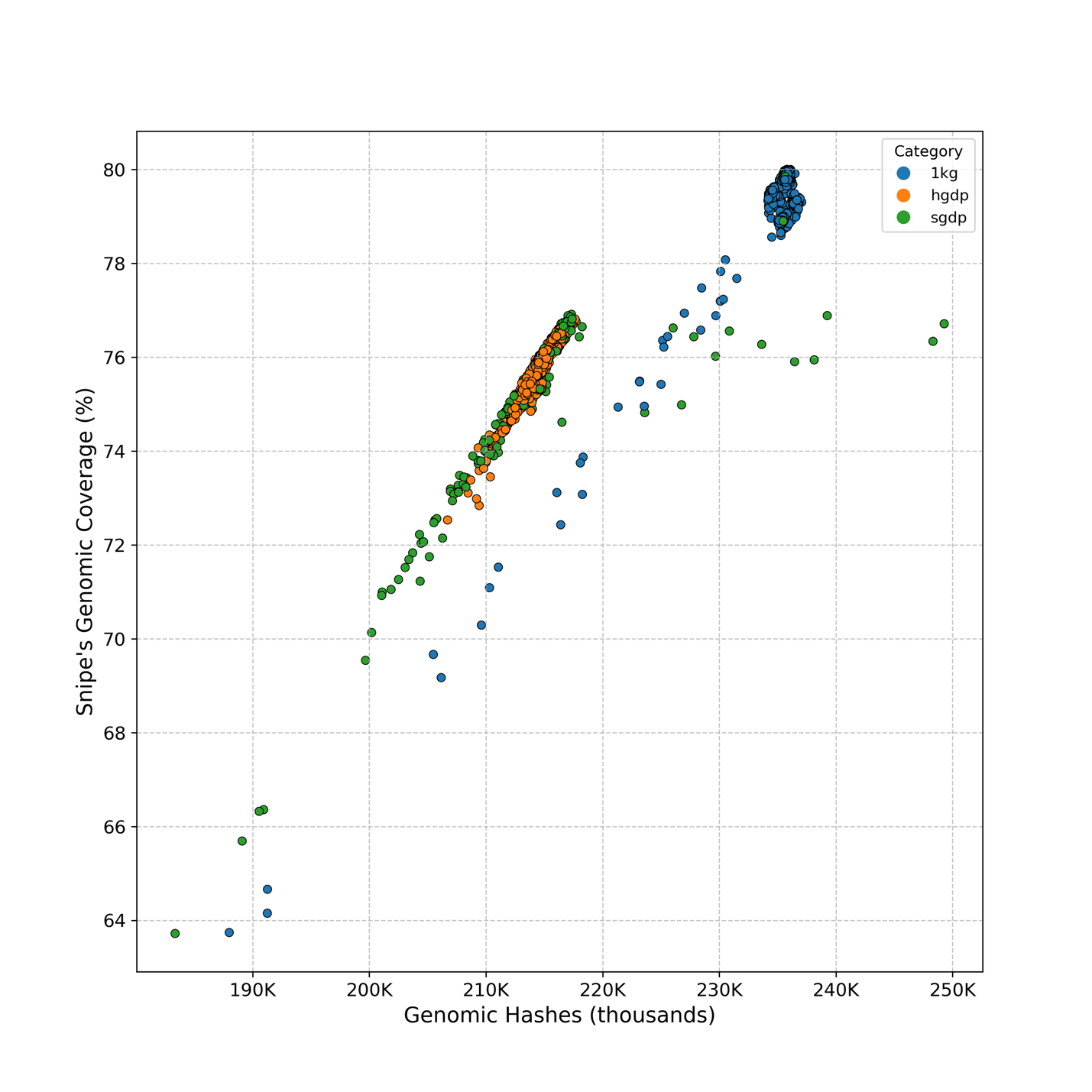

**Supplementary Figure 2: Alignment-free estimation of the human genome coverage in the 1kGP samples**

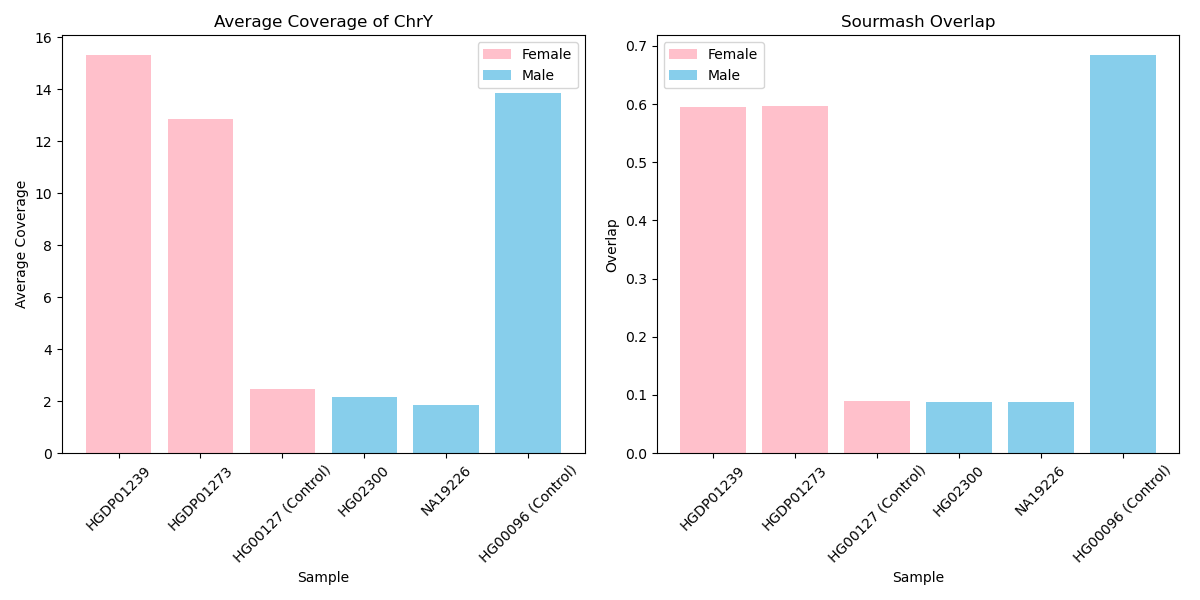

**Supplementary Figure 3:** **Gender discrepancies in four samples of the 1kGP metadata**. The figure shows an agreement between alignment-free (right panel) and alignemnt-based (left panel) gender detetion while disagreeing with the online metadata. On the right, the alignment-free approach identify the gender by calculating the containment between the sample’s signature and the signature of the reference chromosome Y. These four samples showed unexpected deviation from the mean of its gender containment ratio (0.65 with SD=0.039 and 0.08 with SD=0.003 for males and females respectively). On the left, average sequencing coverage of chrY was calculated from the cram files. The color of the bars represents the sex as provided in the metadata, with blue bars denoting male and pink bars denoting female. Ideally, blue bars should be larger than pink bars; however, inconsistencies arise due to errors in the metadata.

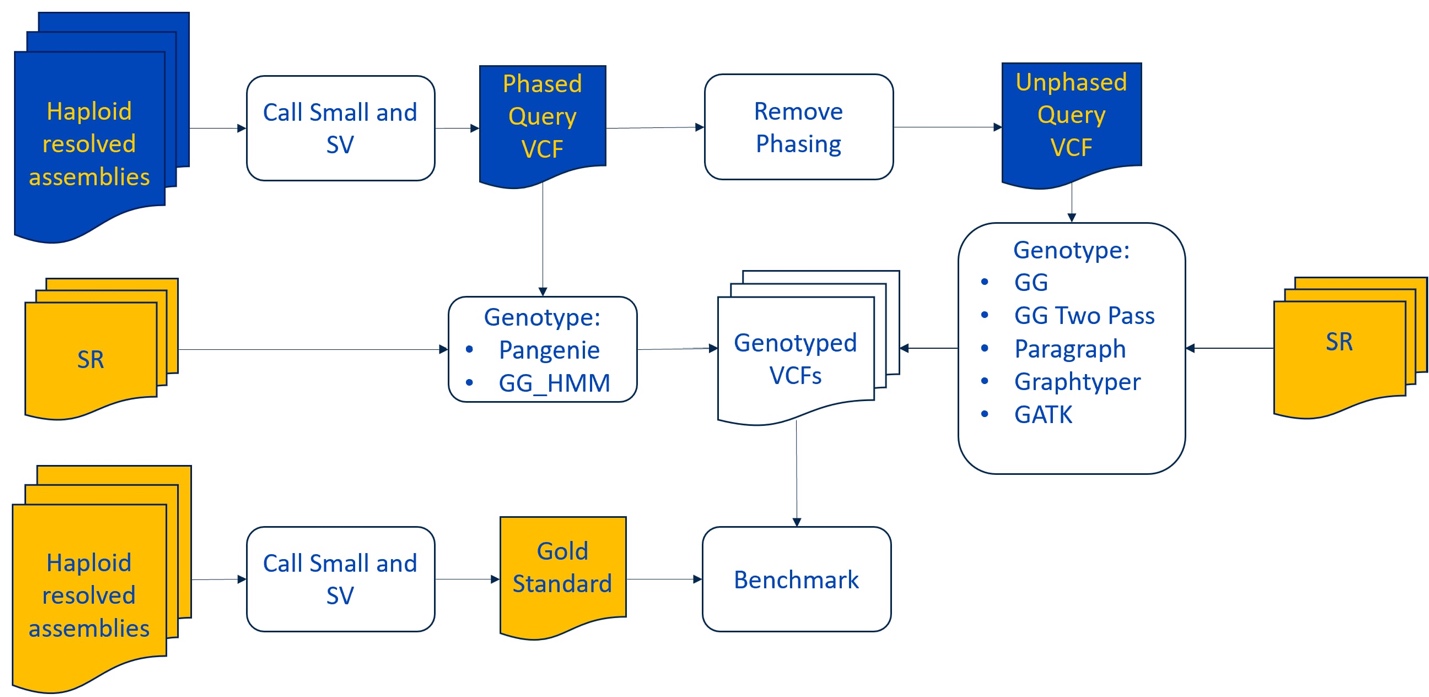

**Supplementary Figure 4.** **Benchmark Genotyping Accuracy Workflow:** The figure represents the workflow for the benchmarking experiment. The gold color represents samples: HG00731 and the blue represents the NA12878 sample.

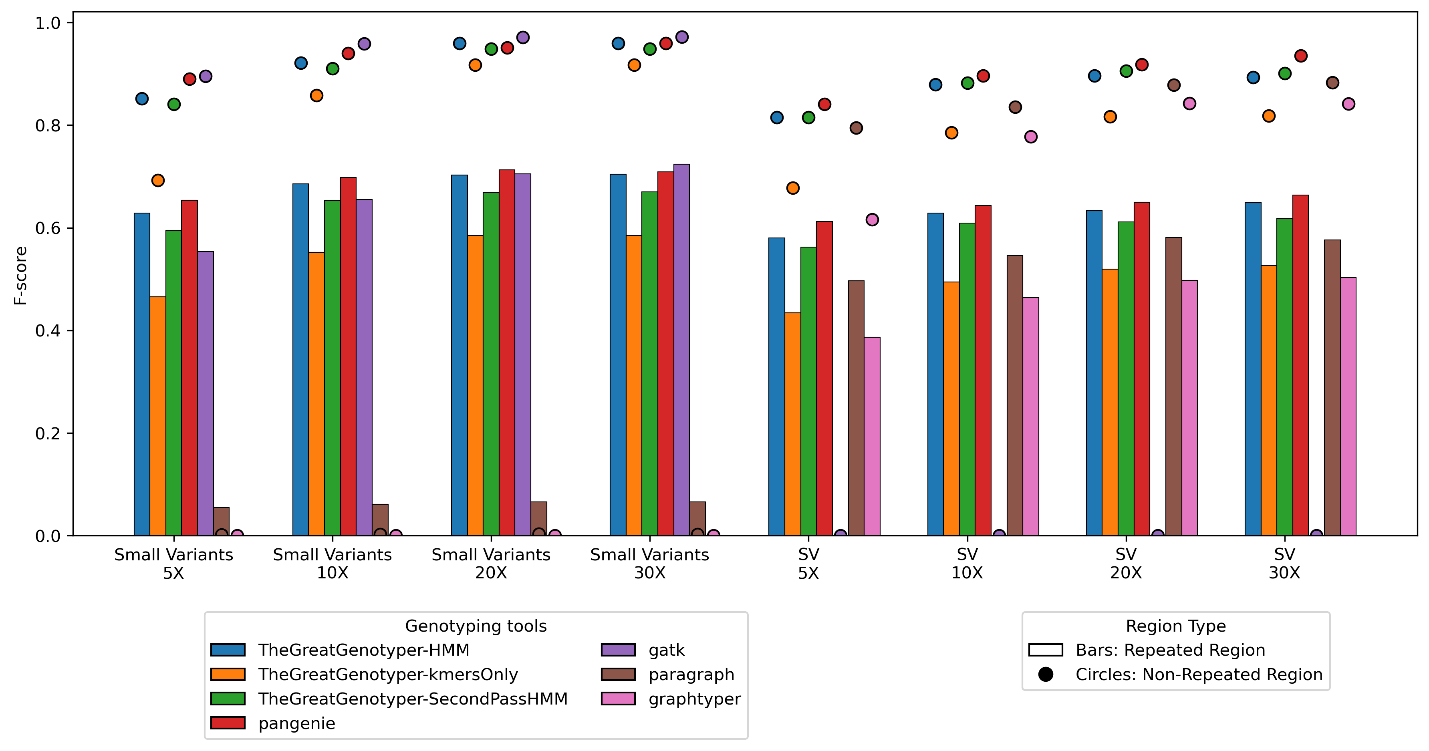

**Supplementary Figure 5.** **Coverage effect on genotyping F-score**: The figure illustrates the effect of coverage on the F-scores of different genotyping methods, differentiating between small variants (under 50 bp) and SV (above 50 bp). F-scores for variants located in repeated regions are shown as bars, while those in non-repeated regions are shown as circles. Additionally, variants are categorized based on the complexity of their genomic location.

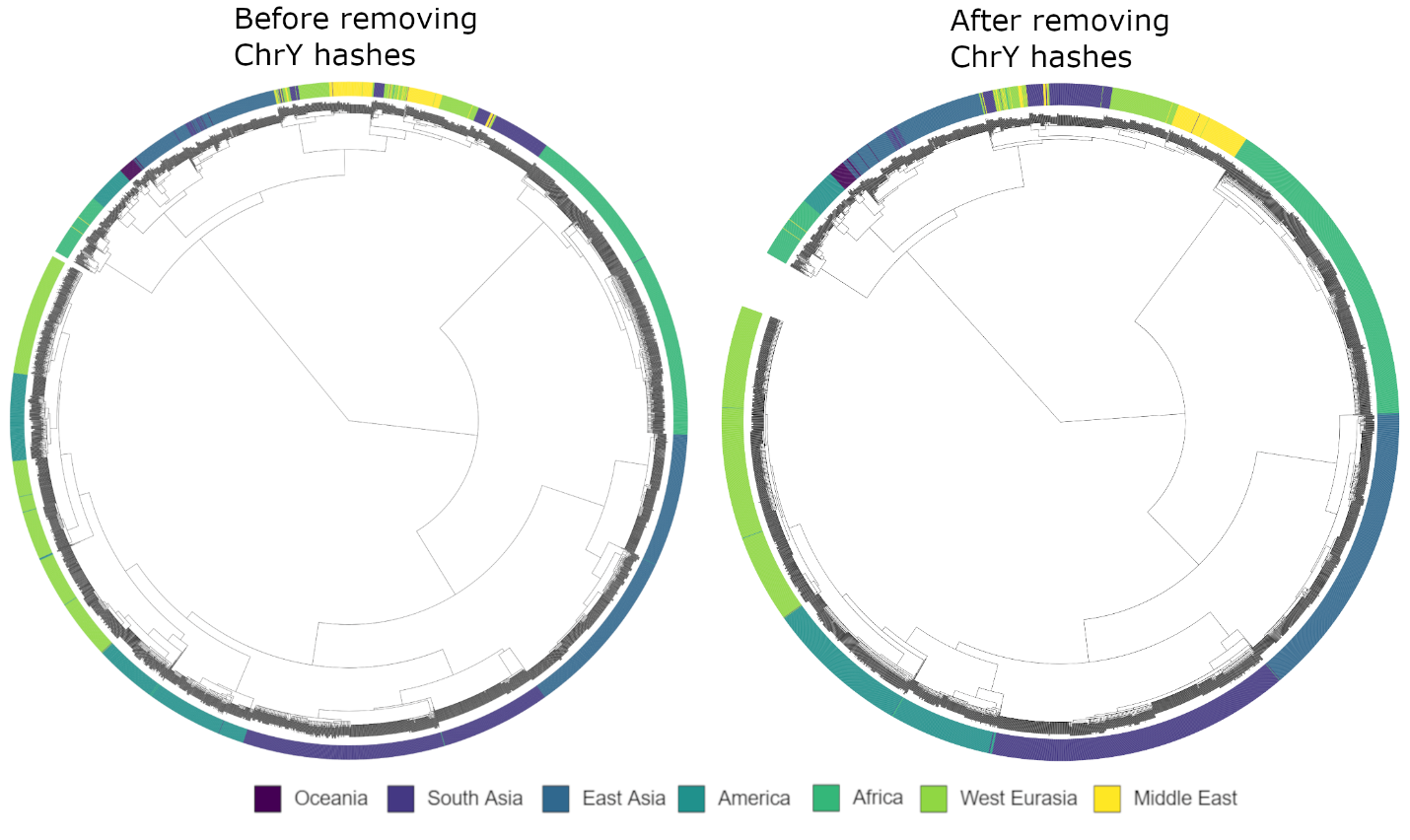

**Supplementary Figure 6. Addressing chrY Bias in Clustering**: The figure depicts two attempts to create a dendrogram for the 4271 samples. The outer circles consist of fine lines, each representing a sample, with the color of the lines signifying the population of the sample as per the metadata. The dendrogram is displayed within these circles, delineating the clusters. Excluding chrY hashes yields more homogeneous clusters, as illustrated in the left dendrogram.

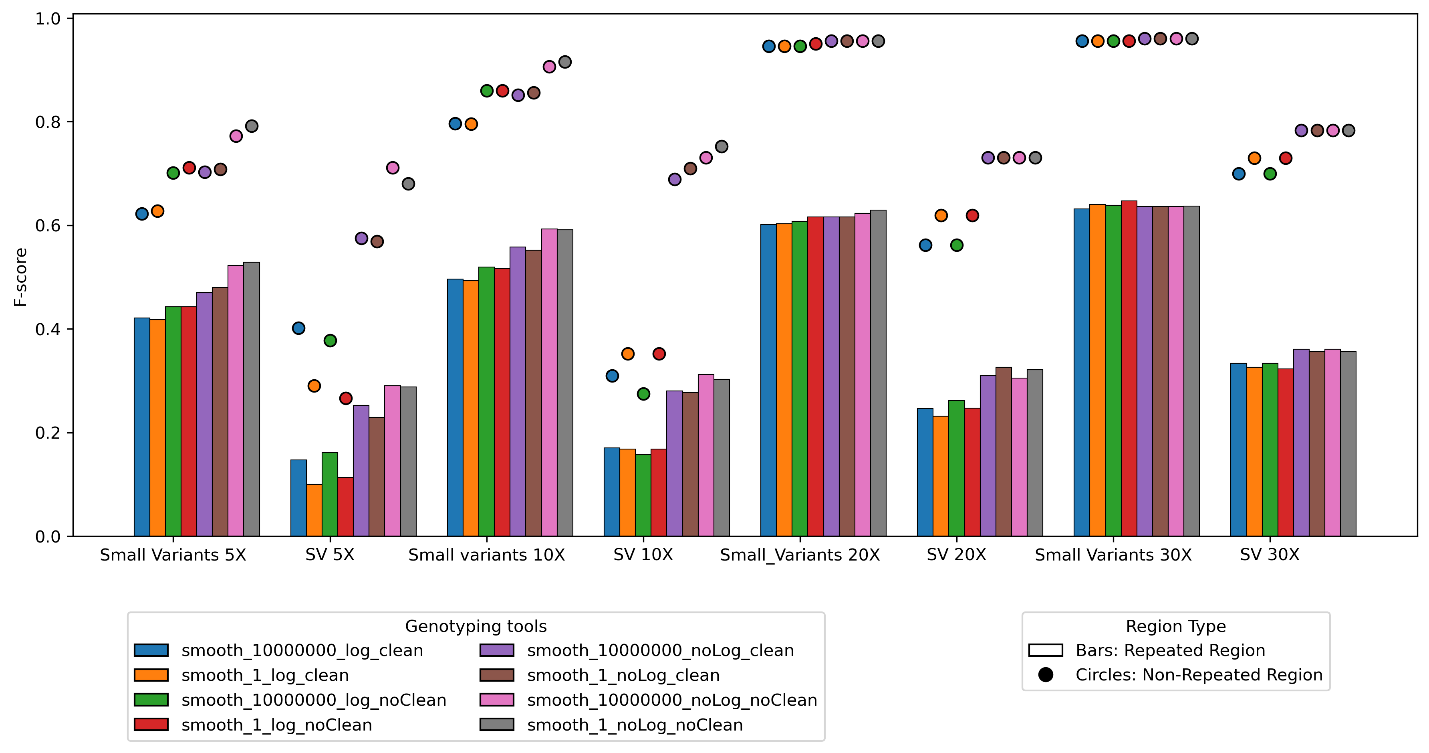

**Supplementary Figure 7.** **F-score comparison between The different parameters of indexing**: Metagraph's preprocessing techniques influence the genotyping f-score. These techniques include *Smoothing Counts*, where only the average kmer count per unitig for each sample(smooth10000000) is retained instead of preserving the individual kmer counts(smooth1); *Log Counts*, which involves saving the log of kmer counts to conserve space; and *Clean*, the error cleaning algorithm in the Metagraph documentation(https://metagraph.ethz.ch/static/docs/quick_start.html#graph-cleaning)

| Variant Type | Count |
| --- | --- |
| SNV | 18.5M |
| Indels(<50 bp) | 7.3M |
| Insertions | 1.5M |
| Deletions | 1.8M |
| Complex | 4.1M |
| SV(>=50bp) | 904.7K |
| Insertions | 85.5K |
| Deletions | 23.4K |
| Complex | 795.8K |

**Supplementary Table 1.** **Variant Counts in HPRC Pangenome** The table details the number of variants from the decomposed VCF of the HPRC pangenome stratified by their type.

| Index Name | Smoothed Counts | Log Counts | Cleaned | CCDG size(GB) |
| --- | --- | --- | --- | --- |
| Smooth10000000_log_clean | Y | Y | Y | 1.42 |
| Smooth10000000_log_noClean | Y | Y | N | 1.59 |
| Smooth10000000_noLog_clean | Y | N | Y | 1.62 |
| Smooth10000000_noLog_noClean | Y | N | N | 1.80 |
| Smooth1_log_clean | N | Y | Y | 1.76 |
| Smooth1_log_noClean | N | Y | N | 1.92 |
| Smooth1_noLog_clean | N | N | Y | 3.00 |
| Smooth1_noLog_noClean | N | N | N | 3.30W |

**Supplementary Table 2.** **Impact of Metagraph Preprocessing on CCDG size:** The table demonstrates how preprocessing methods like Smoothing Counts, Log Counts, and the Clean algorithm alter the size of the final CCDG. Smoothing Counts simplifies kmer data to averages per unitig, Log Counts compresses kmer information logarithmic ally, and Clean removes errors.
